# Mind the gap: emergent clinical risk at the interface of two individually safe AI systems in a multilingual ambient scribe

**DOI:** 10.64898/2026.09.22.26363642

**Authors:** Henry Bergman, Vivian Liu, Ben Austin, Rohan Sanghera

## Abstract

**Objectives:** To determine whether single-layer evaluation characterises clinical risk in the final note of a multilingual ambient AI scribe, and where serious errors arise.

**Design:** Two-arm evaluation on one common clinical-risk scale, using a frozen, reference-aligned synthetic corpus.

**Setting:** Scripted consultations spanning 77 languages and five complexity levels, from simple general practice to expert multidisciplinary-team handover.

**Main outcome measures:** Per-session incidence of at least one serious (HIGH or CRITICAL) note-layer error in intrinsic (reference script to note, n=385) and end-to-end (automatic speech recognition (ASR) transcript to note, n=2,302) generation; three LLM raters classified discrepancies using a four-mode taxonomy.

**Results:** Intrinsic note generation had a 3.4% serious-error rate, with no detectable gradient across complexity (1% to 6%; Cochran-Armitage p=0.42) or language resource (high 2%, medium 5%, low 4%). End-to-end serious errors rose steeply with complexity (L1 1% to L4/L5 24%) and language scarcity (high 7%, medium 10%, low 15%; OR 1.54 per tier, 95% CI 1.15-2.07; p=0.004, GEE clustered on language). Of serious in-note errors, 87% were ASR-derived and 7% note-originated; amplification exceeded correction threefold (fate entropy 1.19 bits). Repeated generation showed moderate reproducibility (Fleiss kappa 0.42); word error rate explained 24% of cross-language variance versus ∼1% for transcription risk density.

**Conclusions:** Low serious-error rates in individual layers did not preclude higher end-to-end note risk. Evaluation should therefore include the clinician-facing note, because component-level metrics cannot fully capture risk created or transformed at the transcription-to-generation interface.

**What is already known on this topic:**

- Ambient AI scribes are being deployed globally, and safety evaluation has characterised the transcription layer in isolation, most commonly with word error rate
- Frequency-based transcription metrics such as word error rate do not directly encode clinical consequence, and component-level performance may not represent the risk of the final generated note
- Large language models produce clinically relevant output stochastically, so identical prompts can yield materially different content

**What this study adds:**

- Note generation from a perfect transcript had a low observed serious-error rate and no detectable complexity or language-resource gradient, whereas the end-to-end system showed steep gradients on both axes.
- Serious note-layer errors were predominantly ASR-derived, but note generation transformed upstream errors variably, amplifying serious errors about three times more often than it corrected them
- Transcription-layer metrics had limited predictive value for note-layer risk after accounting for complexity and language-level dependence; word error rate explained approximately one-quarter of cross-language variance

**How this study might affect research, practice or policy:**

- Evaluation and procurement of ambient scribes should include end-to-end assessment of the clinician-facing note, with results stratified by consultation complexity and language where relevant
- A three-surface root-cause alignment using a four-mode root-cause taxonomy (note-originated, propagated-ASR, amplified-ASR and silent-correction) can attribute documentation errors to their point of origin or transformation
- Language-related performance should remain a surveillance target because disparities can emerge after components are composed, even when corresponding gradients are not detectable in an individual layer

## Introduction

Ambient AI medical scribes are being deployed globally to draft documentation from consultation audio, with early trial evidence of reduced documentation time and clinician burnout,^1^ amid debate over how documentation quality should be evaluated for this device class.^2^ The pipeline has two stages: an ASR model transcribes the audio, and a large language model (LLM) generates a structured note. Safety evaluation has largely characterised the transcription layer alone, most often via word error rate (WER),^3,4^ assuming an accurate transcript yields a safe note and component accuracy certifies system-level safety.

Component-level evaluation can fail for two reasons. First, error frequency and clinical consequence are distinct constructs: frequency-based metrics correlate imperfectly with clinically relevant quality in generated text^5^, and only a minority of speech-recognition errors are clinically significant or persist to the signed record^6,7^; a component metric used to support a safety claim is a surrogate that must be empirically related to the outcome it represents^8,9^. Second, note generation introduces its own failure modes: a generative model can omit or fabricate content^10^, or transform an upstream error into a more or less consequential statement, and identical inputs can yield materially different outputs^11^.

In a companion analysis of the same corpus we evaluated the transcription layer alone: across 77 languages, WER was not associated with dangerous transcription-error rate, and in-context transcription risk showed no detectable gradient by language resource.^12^ That analysis stopped at the transcript, yet the artefact a clinician acts upon is the note; note generation may propagate, correct, amplify, omit or introduce content, none inferable from the transcript alone — a systems-safety problem, where serious failures arise from interactions between components adequate in isolation.^13^

Here we ask whether single-layer evaluation can certify the safety of the final note, and characterise where, how and for whom serious documentation errors arise, evaluating note generation in isolation from a perfect transcript and end-to-end from the characterised ASR transcript, on the same risk scale as the transcription layer. We also propose a root-cause taxonomy attributing each error to its originating layer, and derive recommendations for end-to-end evaluation and post-market surveillance.

## Methods Corpus

We used the frozen synthetic corpus from our earlier transcription-layer analysis: five dictation scripts spanning a complexity gradient (L1 simple general practice visit, L2 moderate general practice review, L3 clinical outpatient, L4 dense ward round, L5 expert multidisciplinary-team handover), authored in English and translated into native-script reference transcripts for 77 languages with back-translation review, then frozen; translation rather than independent authorship held clinical content constant. Each reference was rendered to neural multi-speaker text-to-speech under three acoustic conditions, with up to two voices per language, and the production ASR yielded a transcript for every session alongside the fixed reference script.

### Two note-generation arms with a transcription-layer comparator

The transcription layer (reference script to ASR transcript) had been characterised on the same corpus and risk scale, used here as a contextual comparator, not a third arm. In the intrinsic arm, the model received each ground-truth transcript and produced a note (n=385; 77 languages by five complexity levels), isolating errors despite a perfect transcript; in the end-to-end arm, the same model received each frozen ASR transcript (n=2,302). Frozen transcripts, rather than live audio, maintained alignment of reference, transcript and note by construction; one transcription-failure artefact was excluded. Notes were generated in the consultation language through the production endpoint under default settings (figure 1). The model version and configuration are commercially sensitive and withheld, limiting reproducibility; discussed further in the Limitations.

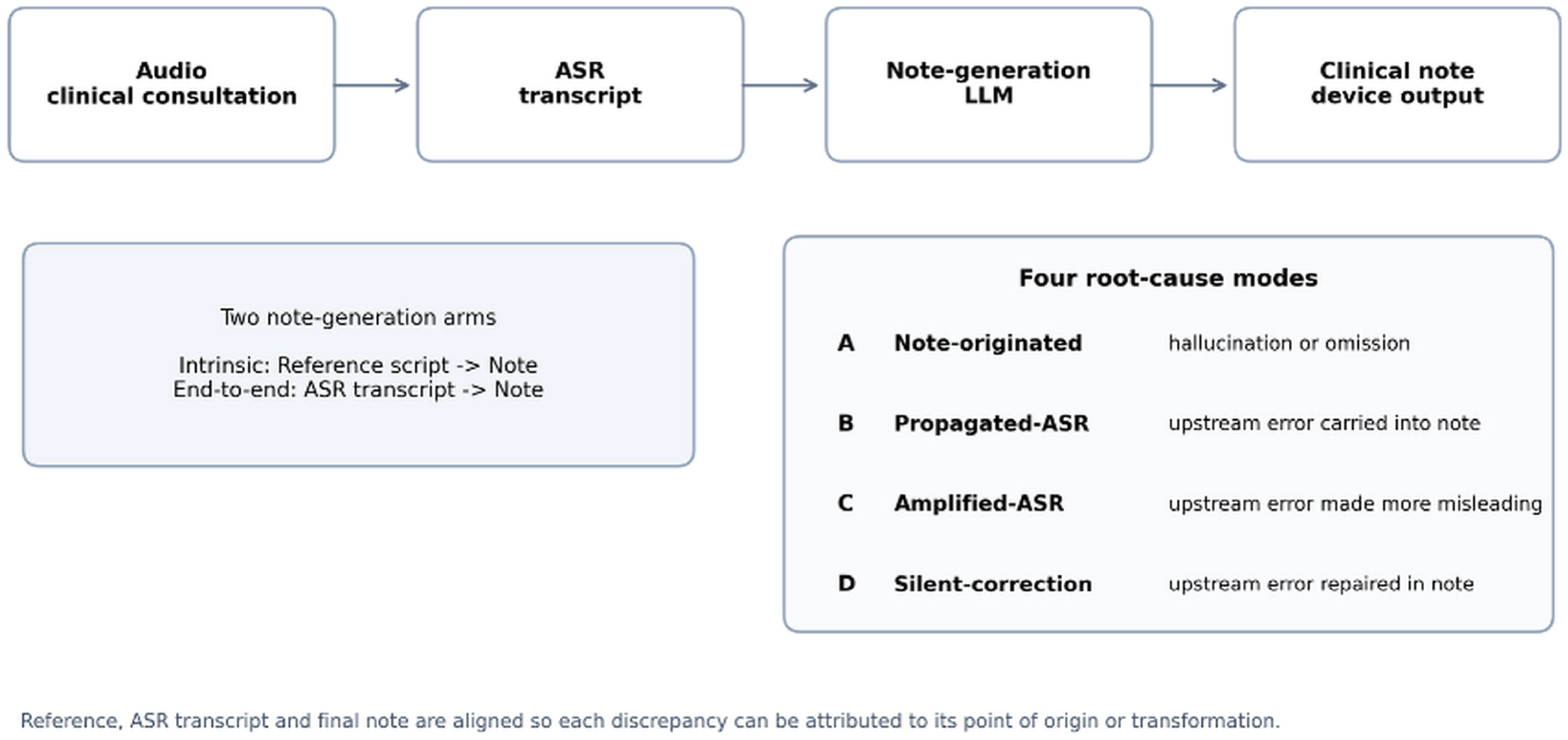

### Error taxonomy

Each discrepancy was classified by comparing reference, ASR transcript and note, with the root-cause mode assigned by three-rater majority vote against an explicit rubric, using four modes: note-originated (A, content introduced or omitted despite correct support in the reference/transcript; subdivided into hallucination and omission), propagated-ASR (B, a transcription error carried faithfully into the note without becoming more misleading, including omissions that remain omitted), amplified-ASR (C, a transcription error transformed into a more misleading statement), and silent-correction (D, repaired using the wider clinical context). The propagated/amplified boundary depended on clinical consequence, with an ASR-grounded mode preferred when attribution was ambiguous unless the note introduced unsupported content, limiting over-attribution to note generation.

### Severity and rater protocol

Each discrepancy was assigned a clinical-risk tier using a Severity x Likelihood score aligned with medical-device and UK digital clinical-safety risk-management practice, including ISO 14971 and DCB0129/DCB0160.^14 15^ Severity (1-5) was the potential consequence if believed; likelihood (1-5) was the probability it would survive review and propagate to harm. Four tiers were defined: LOW (1-4), MEDIUM (5-9), HIGH (10-15) and CRITICAL (16-25), with HIGH or CRITICAL considered serious. Errors were judged in context and in the consultation language, as if a clinician relied on the note (for silent corrections, severity is harm averted); the same instrument was used at the transcription layer for comparison. Three independent LLM raters extracted, classified and graded each note; errors detected by at least two of three were reconciled at median severity. In-language LLM adjudication made evaluation across 77 languages tractable, an acknowledged limitation.

### Reliability and audit

Detection agreement was summarised from the per-error rater-detection count. Severity reliability was assessed by independent blind re-scoring of a stratified 200-error sample by three fresh raters, yielding Fleiss kappa,^16^ quadratic-weighted kappa,^17^ exact and within-one-tier agreement. We separately audited non-genuine flags from reference corruption or benign formatting, removed only cases where the note demonstrably carried the correct value, and re-ran the headline analyses as a sensitivity check.

### Statistical analysis

The primary endpoint was per-session incidence of at least one serious note-layer error.

Risk-tier-weighted densities per 1,000 words were reported for mechanistic detail (band-midpoint weights: LOW 2.5, MEDIUM 7, HIGH 12.5, CRITICAL 20). Performance was decomposed by complexity and language resource tier (taxonomy of Joshi et al.^18^); complexity trend used the Cochran-Armitage test,^19^ and decoupling of transcription- and note-layer risk used Spearman correlation and AUC overall, within strata and by language, with language-cluster bootstrap intervals. ASR-error fate predictability was summarised by Shannon entropy; given the reliability profile, the robust unit was serious vs not serious, with per-tier counts indicative. Analyses used Python (pandas,^20^ scipy,^21^ statsmodels,^22^ pingouin^23^).

### Clustering and dependence

Observations were clustered by language (77) and script (5), with further nesting by voice and noise condition. Between-language contrasts used GEE logistic regression (exchangeable working correlation, sandwich standard errors clustered on language); within-language contrasts additionally used language as the analytic unit. The intracluster correlation for a serious end-to-end note-layer error was 0.077 (design effect 3.24). Because each complexity level maps to exactly one script, a script random effect is not identifiable alongside complexity, so the complexity axis is interpreted as an ordered-script effect; full cluster-respecting sensitivity analyses are in supplementary table S1.

## Results

### Reliability of the rating instrument

Detection agreement increased with risk tier: unanimous detection was 99% for CRITICAL, 96% for HIGH, 84% for MEDIUM and 51% for LOW, the lower LOW-tier figure expected under a heavily skewed marginal, where chance-corrected statistics behave counterintuitively.^24^ Blind independent re-grading gave Fleiss kappa 0.774^25^ and quadratic-weighted kappa 0.906,^17^ with 84% exact and 100% within-one-tier agreement; disagreement never exceeded one adjacent tier and a CRITICAL error was never re-graded below HIGH (supplementary table S2). These figures quantify agreement among LLM raters rather than clinician inter-rater reliability, and were highest in the serious band driving the principal analyses.

### Intrinsic note generation shows low serious-error rates with no detectable gradient

From a perfect transcript, 265 canonical errors arose across 385 notes; 3.4% of notes contained a serious error (12 HIGH, one CRITICAL), with no significant complexity gradient (1% to 6% across L1-L5; Cochran-Armitage z=0.80, p=0.42) or resource gradient (high 2%, medium 5%, low 4%) (supplementary table S3). The L4 peak rests on five of 77 notes, with a wide 95% CI (2.8-14.3%, Wilson) overlapping every stratum; given the single generation per pair and low base rate, this should be read as no detectable gradient rather than evidence of invariance.

### End-to-end note risk rises with complexity and language scarcity

With the real ASR transcript as input, 11.5% of notes (264/2,302) contained a serious note-layer error, rising sharply with complexity (L1 1%, L2 3%, L3 6%, L4 24%, L5 24%) and language scarcity (high 7%, medium 10%, low 15%). The cluster-robust between-language estimate was OR 1.54 per tier step (95% CI 1.15-2.07; p=0.0035, GEE clustered on language), and the gradient held independently within 58 of 60 informative languages (sign test p<0.001), not a pooling artefact — contrasting with the low, non-gradient intrinsic arm, consistent with risk being created or transformed when transcription and note generation are composed end-to-end (figure 2; supplementary table S1).

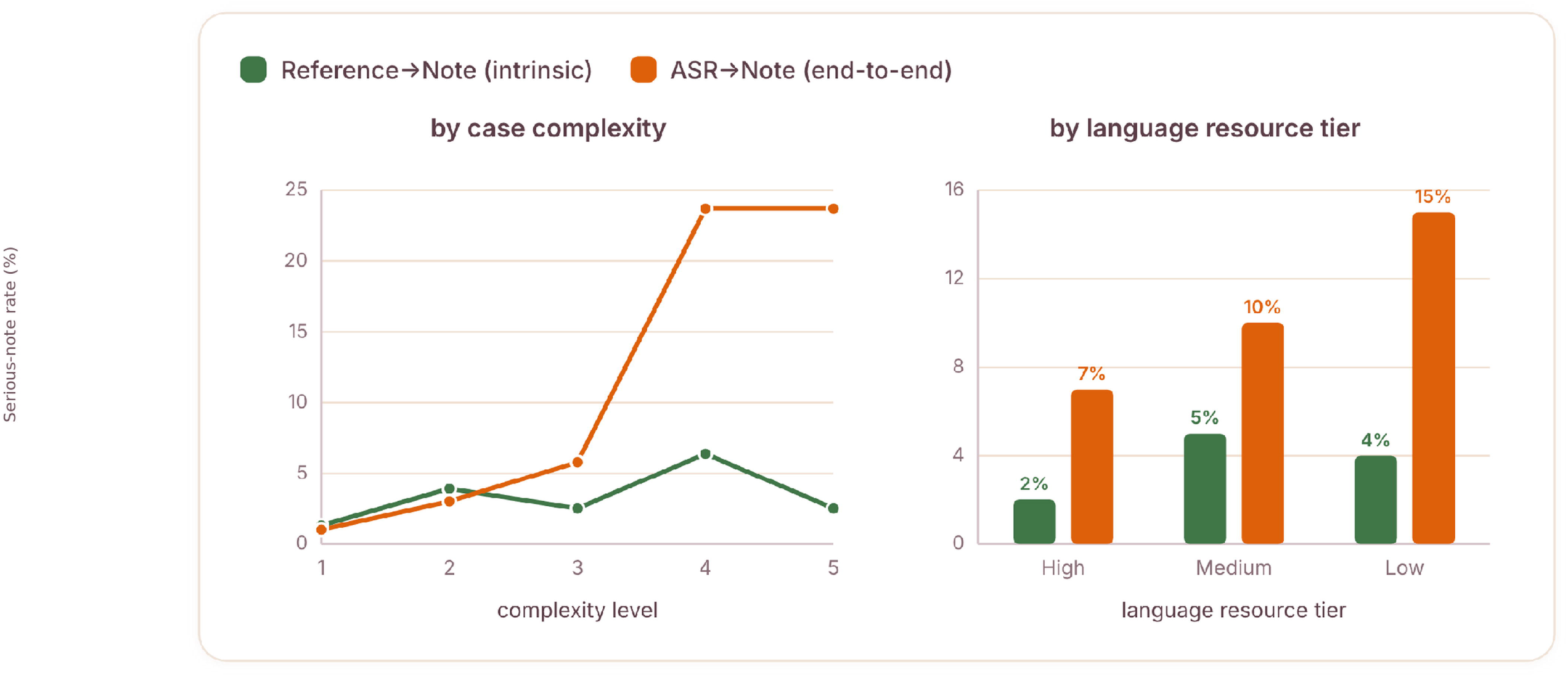

### Most serious errors are ASR-derived and variably transformed

Across all 6,360 canonical errors after audit, the four root-cause modes were propagated-ASR (4,003: 2,839 propagated, 1,164 ASR omissions), note-originated (1,099: 684 hallucinations, 415 omissions), silent-correction (708) and amplified-ASR (550) (table 1; risk-tier distribution in supplementary figure S1). Within the serious-error subset (HIGH or CRITICAL), 87% were ASR-derived and 7% note-originated: most serious errors originated upstream, with a smaller proportion arising de novo during note generation.

**Table 1.** Four-mode root-cause taxonomy and risk-tier distribution, ASR transcript to Note arm.

| A. Four-mode root-cause taxonomy |  |  |  |
| --- | --- | --- | --- |
| Primary mode | Component attribution | All canonical errors, n | Reported subtypes |
| A. Note-originated | Note-generation layer | 1,099 | Hallucination 684; omission 415 |
| B. Propagated-ASR | Transcription layer | 4,003 | Propagated 2,839; ASR omission 1,164 |
| C. Amplified-ASR | Interface | 550 | Upstream error rendered more clinically misleading |
| D. Silent-correction | Note-generation layer (benefit) | 708 | Upstream error repaired in the note |
| **Total** |  | **6,360** |  |
| B. Risk-tier distribution across all canonical errors |  |  |  |
| Risk tier |  | Count |  |
| CRITICAL |  | 153 |  |
| HIGH |  | 1,297 |  |
| MEDIUM |  | 2,495 |  |
| LOW |  | 2,415 |  |

Table 1 reports all canonical errors; supplementary figure S1 shows the four primary modes by risk tier.

The note layer transformed upstream risk variably: among ASR-derived errors with a serious downstream outcome, 274 were amplified and 90 silently corrected, roughly three to one; across all ASR-derived errors, 69% were propagated, 13% amplified and 17% corrected, giving a fate entropy of 1.19 bits. Overall, the note layer contributed 370 serious outcomes through amplification or note-originated error, against 90 averted through silent correction (supplementary figures S2 and S3), robust to the propagated-versus-amplified classification boundary (sensitivity bounds in supplementary table S1).

Serious errors were rarely introduced de novo; more commonly, upstream errors were converted into fluent prose and propagated or amplified.

### The serious-error signal is partly stochastic

To separate structural interface behaviour from generation noise, we regenerated notes three times from the identical frozen ASR transcript for an 80-session, complexity-stratified subsample enriched for L4/L5. Reproducibility of the serious endpoint was only moderate: Fleiss kappa 0.42, with runs unanimous in 68% of sessions (54/80); among 34 sessions serious in any run, only 24% (8/34) were serious in all three, 16 in one run and 10 in two (supplementary table S4) — classification differed across repeated generations in 32% of sampled sessions.

This has two consequences: per-session and per-error estimates, including the 1.19-bit fate entropy, are means over a stochastic process rather than deterministic properties, and non-determinism is clinically relevant since the same input can cross the serious-error threshold on regeneration. As this experiment used a single adjudicator, judge noise is folded into the flips, so 68% concordance is a lower bound on true reproducibility; it was performed only in the end-to-end arm and does not quantify stochastic uncertainty in the intrinsic arm.

### The resource gradient is predominantly ASR-mediated

Decomposed by severity-weighted density per 1,000 words, note-originated serious density rose only mildly with scarcity (high 0.09, medium 0.14, low 0.21), whereas ASR-derived serious density was larger and rose about 2.5-fold (high 1.19, medium 2.11, low 3.03), dominating the gradient (supplementary table S5); silent corrections also increased with scarcity (116, 190, 404) but did not offset it. The end-to-end resource gradient is therefore consistent with an ASR-mediated interface effect, though synthetic speech quality is a potential alternative explanation addressed below.

### Transcription metrics and note-layer risk

At first glance, transcription-layer risk was associated with note-layer risk (AUC 0.88; Spearman rho=0.43, 95% CI 0.38-0.47, language-cluster bootstrap), attenuating within complexity strata (rho 0.14-0.47). At the language level this was not significant: transcription risk density explained ∼1% of cross-language variance (R^2^=0.01, 95% CI 0.00-0.06; rho=+0.19, p=0.09, n=77) (figure 3), against 24% for word error rate (R^2^=0.24, 95% CI 0.05-0.47) (supplementary figure S4) — compatible with 87% of serious errors being ASR-derived, since origin and downstream predictability are different constructs, and upstream risk density remains a weak predictor of which errors become serious.

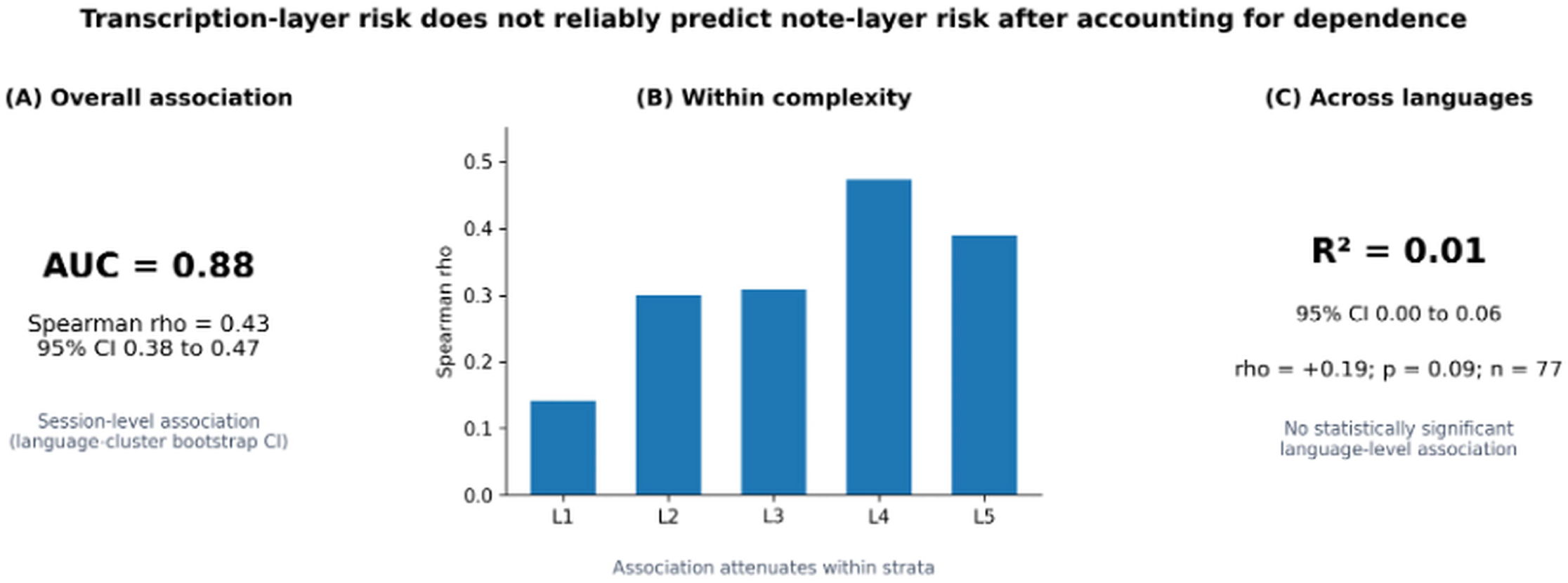

### The clinical character of serious errors

Of 153 CRITICAL errors, 151 were unanimously detected; most were medication or dose errors (99, resembling established look-alike/sound-alike confusions^26^), followed by laboratory-value (18), diagnostic (12, including negation failures^27^) and vital-sign errors (8), concentrated in L4 and L5.

Seven were note-originated and eight were prevented by the note layer, with representative examples, including two ASR-refusal events overridden into confabulated content, given in table 2.

**Table 2.** Adjudicated CRITICAL register: representative serious errors across the three surfaces.

| Reference (truth) | Note (device output) | Class | Severity |
| --- | --- | --- | --- |
| gliclazide | glibenclamide (different sulfonylurea) | Propagated | CRITICAL |
| FOLFOX regimen | FOLFIRI regimen | Propagated | CRITICAL |
| 4-day sore throat (acute) | Chronic 6-month red-flag presentation | Amplified | CRITICAL |
| Chief complaint: sore throat | Chief complaint: knee and back pain | Amplified | CRITICAL |
| Viral illness, no antibiotics | Antibiotics inserted | True-hallucination | CRITICAL |
| gliclazide | sildenafil | True-hallucination | CRITICAL |
| ASR refused: re-dictate | Complete confabulated consultation | True-hallucination | CRITICAL |
| Atrial fibrillation | Atrial restored (ASR said ventricular) | Silent-correction | Prevented |

Class assigned by three-rater majority vote on the six-class taxonomy; severity by median of three independent gradings.

## Discussion

### Principal findings

Across 77 languages, intrinsic note generation showed a low serious-error rate with no detectable complexity or resource gradient, and the transcription layer showed no detectable resource-risk gradient either; the end-to-end system behaved differently, with serious-note errors rising sharply with both complexity and language scarcity. The clinically important signal therefore emerged only once an imperfect transcript was processed by the note-generation model — a failure mode component-level evaluation, including WER, cannot by itself observe, consistent with systems-safety models in which serious failures arise from interactions between components adequate in isolation.^13^

One plausible mechanism is that fluent note generation resolves uncertainty rather than preserving it: on clean input this supports coherent documentation, but on degraded input the same behaviour can convert a low-salience error into a confident clinical statement, helping explain why amplification exceeded correction. Lower-resource languages carry a larger upstream-error burden, creating more opportunities for propagation and amplification, even though intrinsic generation shows no corresponding gradient. Nor is it contradictory that most serious errors were ASR-derived yet language-level risk density explained little downstream variance: origin identifies where an error began, whereas downstream risk depends on how note generation transforms it.

### Strengths and limitations

The principal strength is the two-arm design combined with three-surface alignment on one commensurable risk scale: aligning reference, transcript and note separates intrinsic from end-to-end behaviour, retains the transcription layer as a contextual comparator, and lets each error be attributed to the layer in which it arose.

The corpus is synthetic, providing fixed ground truth and controlled cross-language comparison but not live accent variation, code-switching, disfluency or ASR run-to-run variability — most relevant in lower-resourced settings, where speech-recognition performance is known to vary across speaker populations.^28 29^ Text-to-speech quality may also vary with language resource, so part of the resource gradient could reflect synthetic-audio fidelity rather than a true language-resource effect. Complexity was operationalised as five scripted consultations, one per level, so script identity and complexity are perfectly collinear; the gradient should be read as increasingly complex scripts, not complexity alone. Replication on live audio with multiple scripts per level would address both.

Errors and risk tiers were assigned by three frontier LLM raters from different providers, none the production model under evaluation; this reduces self-evaluation but not independence from shared training data or biases, so reliability measures agreement among automated raters, not clinicians, and tiers should be read as automated estimates. Generation stochasticity is material too: end-to-end reproducibility was only moderate, and the intrinsic arm’s single generation per pair means its null gradients are not proof of invariance. No patient outcome was measured; tiers are simulated, not observed, harm. The system’s version and configuration are commercially sensitive and undisclosed, limiting reproducibility, though the corpus, taxonomy and methodology remain available for replication.

### Comparison with other studies

The low but non-zero rate of omission and fabrication from a clean transcript is consistent with published frameworks for medical-text hallucination and omission^10^ and evidence that generated correspondence can approach junior-clinician quality while retaining characteristic failure modes.^30^ Our repeated-generation result extends evidence that identical prompts yield materially different outputs^11^ by showing non-determinism can flip a safety classification. Decoupling transcription metrics from note-layer harm is consistent with lexical overlap being an inadequate proxy for clinical text quality^5^ and the requirement that surrogate endpoints be validated against outcomes they represent.^8 9^ The clinical plausibility of fluent errors also provides a credible mechanism for automation bias^31^ and helps explain why speech-recognition errors are more readily corrected before signature.^6^

### Implications for verification, validation and surveillance

The device output a clinician relies on is the note, so safety evaluation should be end-to-end on that artefact: a transcript can appear accurate while the note is unsafe, and a poor transcript can still yield an acceptable note, so component testing can misclassify risk in either direction. The two ASR-refusal confabulations imply that note generation should propagate upstream low-confidence signals rather than override them.

Practical implications for pre-market and post-market evaluation, including end-to-end assessment of the clinician-facing note, complexity- and language-stratified reporting and surveillance, and three-surface error attribution, are summarised in supplementary table S6, consistent with evolving European requirements for medical AI devices;^32^ language-related disparity should remain a surveillance target given its independent association with adverse-event harm.^33^

### Future research

Three questions follow directly. First, does the interface behaviour reproduce on live clinical audio, where speaker diversity and ASR variability add error sources absent from synthetic speech? Second, is the serious-versus-not boundary stable under independent clinician adjudication and repeated intrinsic generation? Third, does propagating transcription uncertainty into note generation reduce amplification? The four-mode taxonomy makes these mechanisms experimentally testable.

## Conclusion

In this controlled multilingual corpus, low serious-error rates and no detectable gradients in individual-layer analyses did not preclude substantially higher end-to-end note risk, particularly for more complex scripts and lower-resource languages. Single-layer metrics alone were insufficient to characterise the risk of the clinician-facing note. Evaluation of ambient AI scribes should therefore include end-to-end assessment across the range of languages and consultation complexity in which the system is intended to operate.

## Supporting information

DECIDE-AD Checklist

## Contributors

HB conceived and designed the study, developed the methodology and analysis plan, conducted the analysis, and drafted the manuscript. VL contributed to the methodology and validation, critically revised the manuscript, and contributed to its review and editing. BA contributed to the methodology, regulatory framing, validation, and risk-grading framework, and critically revised the manuscript. RS developed and implemented the software and analysis pipeline, contributed to the investigation, data curation, validation and visualisation, and reviewed and edited the manuscript. All authors approved the final version. HB is the guarantor.

## Competing interests

All authors are employees of Heidi Health, which develops the ambient AI scribe evaluated in this study and funded the work.

### Patient consent for publication

Not required. No patients or patient data were involved.

### Ethics approval

Not required. No human participants and no patient data were involved. All audio was synthesised from author-written scripts containing no patient-identifiable information.

### Patient and public involvement

Patients and the public were not involved in the design, conduct, reporting or dissemination plans of this study. The study is a synthetic device evaluation with no participants.

## Data availability statement

Data supporting the findings of this study are reported in the article and accompanying Supplementary Materials. Additional study materials are provided where appropriate to support interpretation and reproducibility. The underlying evaluation datasets, source materials, and analysis or evaluation code may contain proprietary or commercially sensitive information and are not publicly available. Enquiries regarding access to additional materials may be directed to the corresponding author and will be considered subject to applicable confidentiality, intellectual-property, data-governance, and commercial requirements.

