## Supplementary file for "Mind the gap: emergent clinical risk at the interface of two individually safe AI systems in a multilingual ambient scribe"

**Contents**


| Item | Title | Cited in main text at |
|---|---|---|
| Table S1 | Cluster-respecting sensitivity analyses for complexity, language-resource and decoupling results | Results, cluster-robustness sensitivity analyses |
| Table S2 | Reliability: detection unanimity by tier and blind re-grading agreement | Results, reliability |
| Table S3 | Intrinsic note generation (Reference to Note): serious-note rate by complexity and resource tier | Results, intrinsic arm |
| Table S4 | Generation stochasticity: 80 sessions regenerated three times from an identical transcript | Results, stochasticity |
| Table S5 | Resource decomposition: serious risk-tier-weighted density per 1,000 words | Results, resource gradient |
| Table S6 | Practical implications for pre-market and post-market evaluation | Discussion, implications for verification, validation and surveillance |
| Figure S1 | Four-mode root-cause taxonomy by risk tier | Results, taxonomy |
| Figure S2 | Risk waterfall: the note layer corrects some upstream risk but is net additive | Results, transformation of upstream risk |
| Figure S3 | ASR-error fate by risk tier: per-error fate entropy 1.19 bits | Results, transformation of upstream risk |
| Figure S4 | Single-layer component metrics explain limited cross-language variation in note-layer risk | Results, certification |


**Table S1. Cluster-respecting sensitivity analyses**

Each principal trend or association was re-estimated using methods that respect language-level dependence. The end-to-end language intracluster correlation was 0.077 (design effect 3.24). Complexity is a within-language factor; resource tier is a between-language factor, so clustering chiefly affects the resource-gradient inference.

A. Complexity gradient

| Arm | Published / naive | GEE, cluster=language | Language as unit | Two-stage random effects |
|---|---|---|---|---|
| End-to-end | z=+14.03; p=1.0x10^-44 | OR/level 2.16 (95% CI 1.93-2.41); p=1.6x10^-42 | 58/60 informative languages tau>0; median tau=+0.30; exact p=3.2x10^-15 | Pooled OR (L4/5 vs L1/2) 5.48 (95% CI 3.92-7.67); p=2.8x10^-23; tau^2=0; I^2=0% |
| Intrinsic | z=+0.80; p=0.425 | OR/level 1.18 (95% CI 0.86-1.60); p=0.304 | 6/10 informative languages tau>0; p=0.75 | Pooled OR 1.08 (95% CI 0.67-1.74); p=0.752; tau^2=0; I^2=0% |

B. Language-resource gradient

| Arm | Published / naive | GEE, cluster=language | Jonckheere-Terpstra (77 languages) | Language-level binomial GLM (HC1) |
|---|---|---|---|---|
| End-to-end | z=+5.17; p=2.4x10^-7 | OR/step 1.54 (95% CI 1.15-2.07); p=0.0035 | z=+3.06; p=0.0022 | OR/step 1.54 (95% CI 1.15-2.06); p=0.0036 |
| Intrinsic | z=+0.73; p=0.468 | OR/step 1.29 (95% CI 0.67-2.47); p=0.442 | z=+0.33; p=0.745 | OR/step 1.29 (95% CI 0.67-2.47); p=0.442 |

Language-level serious-note rates (n=77 languages): high resource 7.3% (22 languages), medium resource 9.9% (22), low resource 15.4% (33). Kruskal-Wallis p=0.0087; Mann-Whitney low versus high p=0.0027.

### C. Decoupling and correlation analyses

| Analysis | Published | Cluster-corrected |
|---|---|---|
| Session-level rho: transcription risk density vs serious note-layer error | rho=0.43 | rho=0.425; language-cluster bootstrap 95% CI 0.381-0.469 |
| Within-stratum rho, L1 to L5 | 0.14-0.47 | 0.141 / 0.299 / 0.309 / 0.473 / 0.390; cluster-bootstrap CIs all exclude 0 |
| WER vs per-language serious-note rate, R^2 | 0.23 | R^2=0.235; bootstrap 95% CI 0.049-0.466; rho=+0.39, p=0.0004; n=77 |
| Transcription risk density vs per-language serious-note rate, R^2 | ~0.01 | R^2=0.009; bootstrap 95% CI 0.000-0.062; rho=+0.19, p=0.094; n=77 |

Interpretation: all principal conclusions were unchanged. The resource-gradient p value was the only headline inference materially affected by clustering, moving from 2.4x10^-7 to 0.0035. Complexity is perfectly collinear with script identity (one script per level), so no statistical model can separate those effects without replication using multiple scripts per level. The two quantities most sensitive to the propagated-versus-amplified boundary were explicitly bounded. Reclassifying every serious amplification as propagated reduces the note-layer-attributable share from 25% to 7% and removes amplification; the opposite extreme raises the share to 86% and the amplification-to-correction ratio to 12.8 to 1. The reported estimates therefore lie towards the lower end of the possible note-layer-attributable range, and the conclusion that amplification exceeds correction is robust to the classification boundary.

### Table S2. Reliability of the automated rating instrument

| Severity tier | Unanimous (3/3) detection | Blind re-grade, exact | Blind re-grade, within one tier |
|---|---|---|---|
| CRITICAL | 99% | 84% overall | 100% overall |
| HIGH | 96% | | |
| MEDIUM | 84% | | |
| LOW | 51% | | |

Blind re-grading of a stratified 200-error sample (50 per tier) by three fresh frontier-model raters from different external providers, none used in the production system under evaluation: Fleiss kappa 0.774, quadratic-weighted kappa 0.906. Disagreement never exceeded one adjacent tier and a CRITICAL error was never re-graded below HIGH. These figures quantify agreement among independent LLM raters; clinician inter-rater reliability on this instrument was not measured.

### Table S3. Intrinsic note generation (Reference to Note)

| Stratum | L1 | L2 | L3 | L4 | L5 | Overall |
|---|---|---|---|---|---|---|
| Serious-note rate by complexity | 1% | 4% | 3% | 6% | 3% | 3.4% |

| Resource tier | High | Medium | Low | Serious errors | Canonical errors |
|---|---|---|---|---|---|
| Serious-note rate by resource | 2% | 5% | 4% | 12 HIGH, 1 CRITICAL | 265 across 385 notes |

Serious-note rate is the proportion of notes containing at least one HIGH or CRITICAL error. Cochran-Armitage test for trend across complexity z=0.80, p=0.42. The apparent L4 peak rests on five of 77 notes (95% CI 2.8% to 14.3%, Wilson), overlapping every other stratum. Cluster-respecting sensitivity analyses are reported in Table S1 and leave the intrinsic-arm conclusions unchanged.

#### Table S4. Generation stochasticity

| Metric | Value |
|---|---|
| Per-run serious-error rate (L4/L5-enriched subsample) | 25% |
| Three-run concordance (all runs agree serious versus not) | 68% (54/80) |
| Fleiss kappa across the three runs, serious endpoint | 0.42 (moderate) |
| Of sessions serious in at least one run, serious in all three | 24% (8/34) |
| Serious in exactly one, two or three runs | 16 / 10 / 8 |

Eighty sessions, complexity-stratified and enriched for L4 and L5, regenerated three times from the identical frozen ASR transcript. Because the experiment used a single adjudicator, judge noise is folded into the observed flips, so 68% concordance is a lower bound on true generation reproducibility.

#### Table S5. Resource decomposition: serious risk-tier-weighted density

| Measure | High resource | Medium resource | Low resource |
|---|---|---|---|
| Note-originated serious density per 1,000 words | 0.09 | 0.14 | 0.21 |
| ASR-derived serious density per 1,000 words | 1.19 | 2.11 | 3.03 |
| Silent corrections, count | 116 | 190 | 404 |
| Serious-note rate | 7% | 10% | 15% |

Risk-tier-weighted density uses band-midpoint weights (LOW 2.5, MEDIUM 7, HIGH 12.5, CRITICAL 20). ASR-derived density rises about 2.5-fold across the resource gradient and dominates it, while note-originated density rises only mildly.

#### Table S6. Practical implications for pre-market and post-market evaluation

| Phases | Practical implications |
|---|---|
| Pre-market and ongoing evaluation | Evaluate the clinician-facing note end-to-end; report per-session serious-note incidence; stratify by consultation complexity and language-resource tier; use three-surface alignment to attribute errors as well as count them; avoid treating word error rate or any other single-layer metric as a certification endpoint; and maintain frozen, reference-aligned corpora for regression testing across model and prompt updates. |
| Post-market surveillance | Consistent with evolving requirements for medical AI devices in Europe, monitor serious-note incidence by language and consultation complexity alongside amplification, correction and low-confidence or refusal events. Periodic clinician review of the serious tail and repeated generation on sampled cases would help distinguish device behaviour from evaluator and generation variability. Equity should remain a surveillance target, given the independent association between language barriers and adverse-event harm. |

Implications for verification, validation and surveillance arising from the three-surface, end-to-end evaluation, summarised for pre-market assessment and post-market surveillance.

#### Figure S1. Taxonomy frequency by severity tier

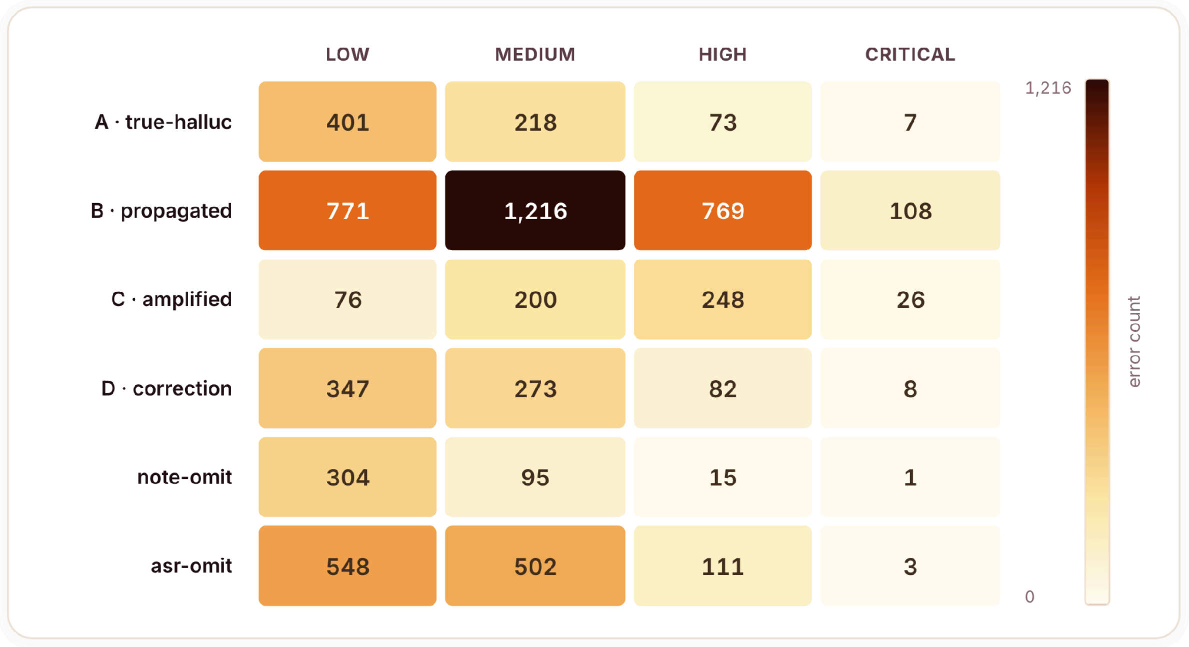


Error counts across all six taxonomy classes and four risk tiers. The original descriptive subtypes have been aggregated as follows: note-originated combines hallucination and note omission; propagated-ASR combines propagated transcription errors and ASR omissions. Propagated-ASR dominates the volume and is concentrated at MEDIUM and HIGH risk.

#### Figure S2. Risk waterfall

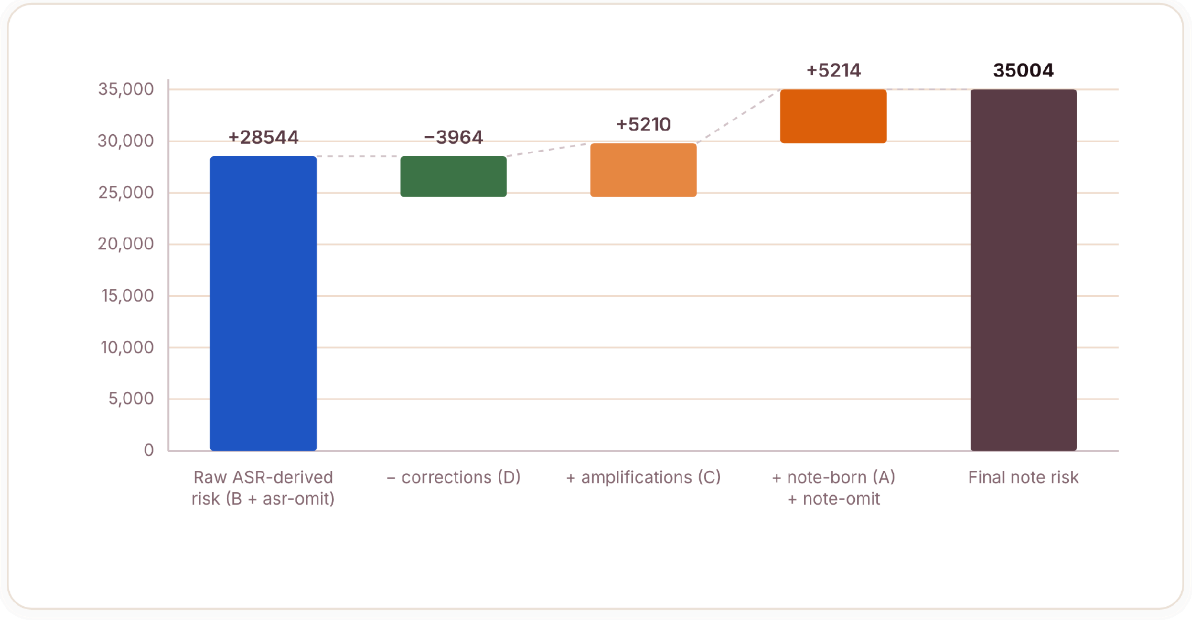


The note layer corrects some upstream risk but is net additive. Severity-weighted risk, summed across MHRA-aligned weights. Raw ASR-derived risk is reduced by silent corrections (-3,964) but more than offset by amplifications (+5,210) and by note-originated errors together with note-omissions (+5,214), giving a net-additive final note risk of 35,004.

#### Figure S3. ASR-error fate by risk tier

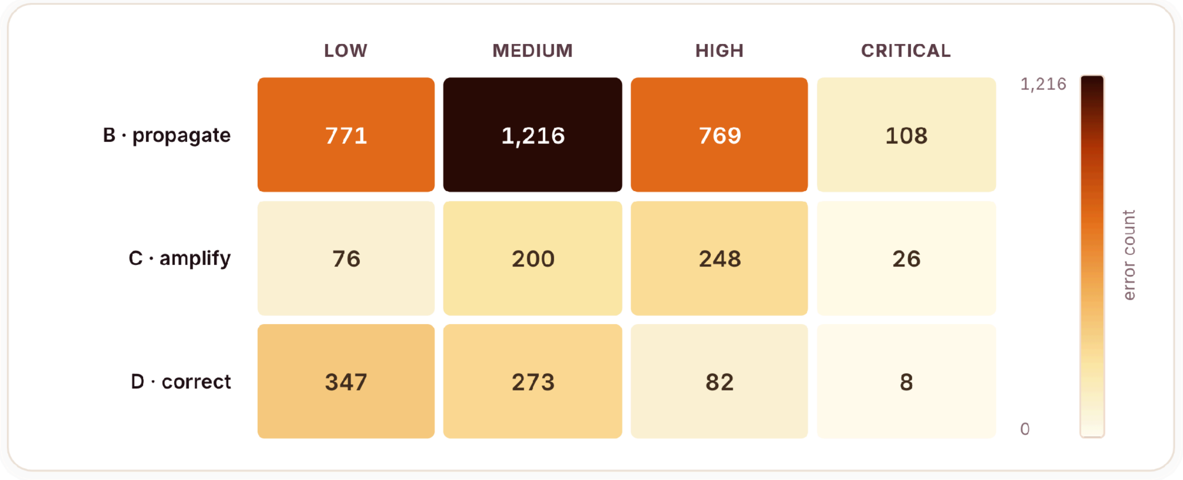


Per-error fate entropy is 1.19 bits, so the fate of an individual upstream error showed substantial uncertainty. Counts of ASR errors by fate (propagated, amplified, corrected) and severity tier. Propagation dominates, but amplification and correction occur across all severities, and there is no severity at which an upstream error's fate becomes predictable. Part of this unpredictability is generation stochasticity (Table S4).

### Figure S4. Single-layer component metrics explain limited cross-language variation in note-layer risk

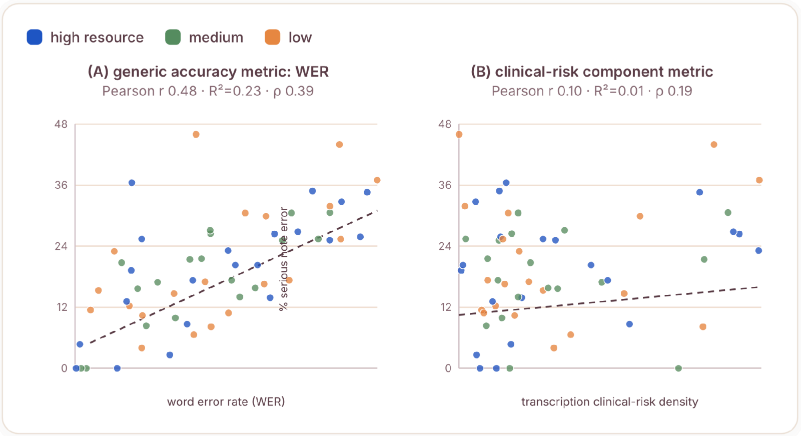


Neither a generic accuracy metric nor a transcription-layer clinical-risk metric predicts per-language note-layer risk. (A) Word error rate explained approximately 24% of cross-language variance in serious note-layer error ($R^2$=0.24, 95% CI 0.05 to 0.47). (B) Transcription clinical-risk density explained approximately 1% ($R^2$=0.01, 95% CI 0.00 to 0.06; rho=+0.19, p=0.09; n=77).
